# A scenario modeling pipeline for COVID-19 emergency planning

**DOI:** 10.1101/2020.06.11.20127894

**Authors:** Joseph C. Lemaitre, Kyra H. Grantz, Joshua Kaminsky, Hannah R. Meredith, Shaun A. Truelove, Stephen A. Lauer, Lindsay T. Keegan, Sam Shah, Josh Wills, Kathryn Kaminsky, Javier Perez-Saez, Justin Lessler, Elizabeth C. Lee

## Abstract

Coronavirus disease 2019 (COVID-19) has caused strain on health systems worldwide due to its high mortality rate and the large portion of cases requiring critical care and mechanical ventilation. During these uncertain times, public health decision makers, from city health departments to federal agencies, sought the use of epidemiological models for decision support in allocating resources, developing non-pharmaceutical interventions, and characterizing the dynamics of COVID-19 in their jurisdictions. In response, we developed a flexible scenario modeling pipeline that could quickly tailor models for decision makers seeking to compare projections of epidemic trajectories and healthcare impacts from multiple intervention scenarios in different locations. Here, we present the components and configurable features of the COVID Scenario Pipeline, with a vignette detailing its current use. We also present model limitations and active areas of development to meet ever-changing decision maker needs.

## Introduction

In late 2019, the virus responsible for coronavirus disease 2019 (COVID-19) was detected in Wuhan, China [1]. Since its emergence, SARS-CoV-2 has spread rapidly, causing significant morbidity and mortality; prompting the World Health Organization to declare a pandemic on 11 March 2020 [2]. In addition to its significant individual health impacts, COVID-19 has put considerable strain on health systems, as a large fraction of cases require mechanical ventilation or critical care [3]. In every stage of the pandemic thus far, there has been a need for flexible decision support tools that can be used to model and compare critical planning scenarios.

Epidemiological models have played an important role in shaping public health policy and interventions throughout the pandemic. The methods used have ranged widely -- from agent-based modeling approaches that simulate the global movement of individuals and their contacts in household, workplace, and leisure settings [4], to population-level models that incorporate features like age-specific transmission, asymptomatic and presymptomatic transmission, and metapopulation structure [5-7], to curve fitting approaches that use data from early in the COVID-19 pandemic to project future burden [8]. Likewise, the goals of these models have varied widely, from assessing importation risk, estimating the fraction of cases attributable to transmission from unobserved infections, projecting the impact of non-pharmaceutical interventions that target different populations, and forecasting the needs of the healthcare system.

Within this space, there was a need for a modeling pipeline that could provide flexible but sophisticated epidemiological models to decision makers who needed to plan and compare specific interventions. Here, we detail our scenario modeling pipeline, a modular framework that projects epidemic trajectories and health care impacts under different suites of interventions in order to aid in scenario planning. The flexibility of our approach has allowed us to provide rapid support to multiple organizations at the same time, while customizing our models to situation-specific questions and data. This framework has been used to provide tailored estimates of the relative impacts of different scenarios of disease transmission, severity, and control, thus guiding intervention policies in several states, countries, and humanitarian aid settings.

## Methods

### Pipeline at a Glance

The pipeline consists of multiple modular components designed to run in sequence to produce results and reports focused on policy relevant outcomes (Figure 1). While the pipeline was developed to be extended, the current core components are (1) epidemic seeding, (2) the transmission model, (3) health outcome generation engine, and (4) report generation.

**Figure 1.**
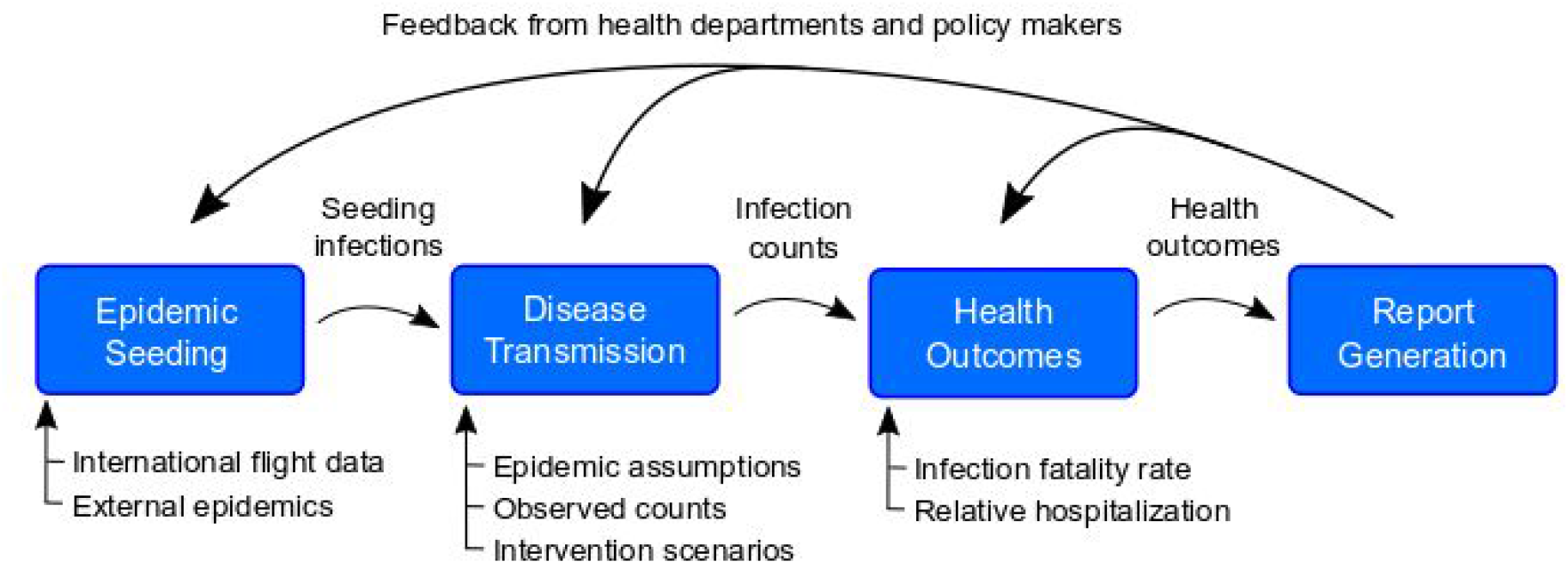
Overview of the pipeline. The pipeline has four modules, each with specific inputs that can be specified by the user. First, we identify when and in which model locations epidemics are seeded using an air importation model or confirmed case data. Second, the epidemic seeding events are used to initiate the disease transmission model, which is informed by our epidemiological assumptions and intervention scenarios. The disease transmission model produces daily incident infection counts and infection prevalence. Next, we calculate health outcomes like hospitalizations and ICU admissions from these infection counts according to assumptions about health outcome risks and infection fatality ratios. Finally, these health outcomes may be summarized using templates and functions from the report generation component of the pipeline.

These modular components of the pipeline fit together because each is composed of multiple pieces: an input format, an output format, one or more code libraries (where applicable), and a runner script. The standardized input and output formats ensure that components may be switched out according to user preference without impacting other phases. The “library” generally contains the core functions for a pipeline component. The script liaises between the user-defined configuration and the library by choosing which library to use and converting the input format to a format used by the library.

The pipeline runs these components in sequence, according to the specifications outlined in a configuration file. This makes it easy to add or modify a component. To add a component, we specify its input format, and incorporate its dependencies. To modify the implementation of a component, we add or modify functions in the library and function calls in the runner script. When appropriate, entire components may be substituted with data from outside of the pipeline, provided that the data meet the input formats required by the next pipeline phase.

### Module 1: Epidemic seeding and initialization

“Epidemic seeding” refers to how the disease transmission module is initialized with infected individuals. A seeding module must produce one of more seeding files that specifies an added number of incident cases occurring due to “seeding” at particular dates and locations.

The pipeline currently contains two epidemic seeding options: 1) seeding according to first case appearance in data, and 2) seeding according to an air travel importation model.

#### Seeding according to earliest identified cases

This seeding option enables users to seed the model according to COVID-19 case data. It currently supports user-supplied data and download from two commonly used public sources, the Johns Hopkins University Center for Systems Science and Engineering (JHU-CSSE) COVID-19 Dashboard [9] and USAFacts, a US-specific database that collates data from state health departments [10]. Drawing from the user-specified data source, this option identifies the first five days that cases were reported in each modeled location. We assume that confirmed cases were infected a user-specified number of days prior to when they were reported, and that there is a user-specified ratio of infections to confirmed cases. Seed infections are, hence, created in each modeled location on the estimated days of infection for the first five days with reported cases; they are drawn stochastically from a Poisson distribution where the mean is the product of the number of reported cases and the user-specified ratio.

To facilitate the generation of the seeding file for US settings, we provide the “R/scripts/create_seeding.R” script in “HopkinsIDD/COVIDScenarioPipeline,” which pulls data directly from USAFacts.

#### Seeding according to an air importation model

We adapted a previously published model of measles importation to model the rate of COVID-19 importation to specific locations due to air travel [12]. This seeding option, available in the Github repository “HopkinsIDD/covidImportation” [13], uses complete itinerary (origin to final destination) air travel volume data from OAG [14] for all airports in the world, source location populations, and source location incidence data, to inform a model with which absolute counts of importation are estimated on a daily basis into airports [13].

Geographic areas surrounding airports are classified spatially into “airport catchment areas” with a Voronoi tessellation of space in reference to the latitude and longitude coordinates of the airport [15]. When there are multiple airports within close proximity, the user may specify a threshold distance under which airports may be grouped into a single cluster that is defined by its centroid. We then assign a probability of importation to each intersection of a Voronoi tile and an administrative unit boundary. This probability is calculated as the proportion of the airport catchment area population that lives in that intersection, assuming that population is distributed evenly by area. All air importations on a given day are then aggregated to the administrative unit level and seeded into the epidemic model as newly infected individuals.

### Module 2: Transmission model and intervention scenarios

The disease transmission module takes in seeding information and produces an epidemic model output file that contains, at minimum for subsequent module compatibility, daily counts of incident infections, indexed to their time of symptom onset. The currently implemented default transmission module comprises a metapopulation model with stochastic Susceptible-Exposed-Infected-Recovered (SEIR) disease dynamics.

#### Disease dynamics

The core model is a modified SEIR compartmental model where the time in the “Infected” compartment follows an Erlang distribution (i.e., the infected compartment is split into *k* compartments) to produce more realistic infectious periods where the chance of recovery depends on the time since infection [16], and a coefficient (*α*) can be set to help the model approximate non-homogeneous mixing between susceptible and infected individuals and non-exponential growth [17]. Currently *k* is fixed at three compartments.

Transition of individuals between disease compartments is simulated stochastically with binomial random draws:

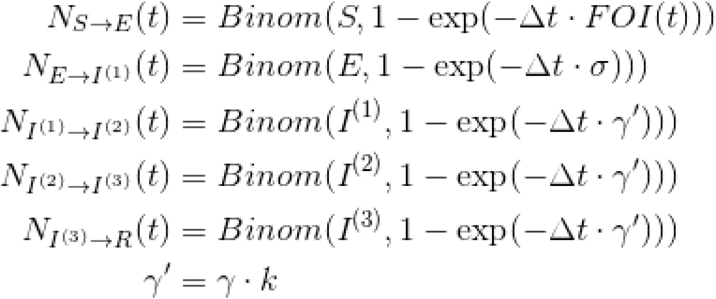

where *S, E, I*^(1)^ *I*^(2)^, *I*^(3)^, and *R* represent the number of individuals in those respective compartments, *FOI*(*t*) is the force of infection from the infected population on the susceptible population, 1/*σ* is the latent period, 1/*γ* is the infectious period, and *k* is the number of *I* compartments. The force of infection, which modulates transition of individuals from the *S* to *E* compartments:

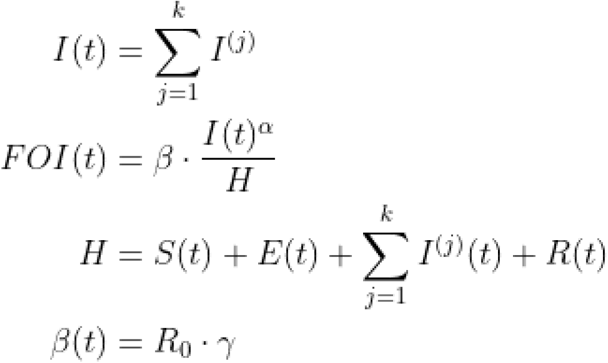

where *H* is the total population, *β* is the daily transmission probability as defined by *R*_0_ and the infectious period, and *a* is the mixing coefficient.

#### Metapopulation dynamics

The model is capable of simulating disease spread in multiple locations jointly according to assumptions about population mobility between individual model locations (e.g., administrative units). The SEIR disease dynamics described above are simulated in each model location with a modification to the force of infection term that accounts for this impact of mobility on disease spread.

The force of infection in a given location *i* is calculated from a combination of local infections and infections in locations that are connected to it according to the mobility matrix, as follows:

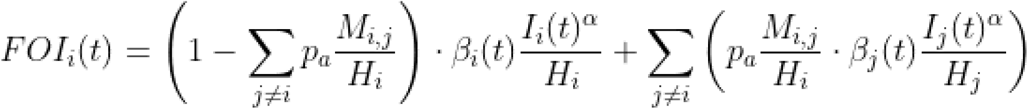

where *M* is a mobility matrix such that *M_i,j_* represents the daily movement of individuals (e.g., commuting) from origin *i* to destination *j, p_a_* is the proportion of time that moving individuals spend away, and *H_i_* is the population of node *i*. The transition of individuals between disease compartments may be modified to index by location *i*, for example:

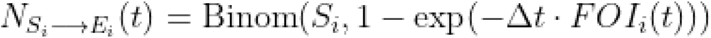

Users may provide a symmetric or asymmetric wide-form mobility matrix for all model locations or a long-form sparse mobility matrix that indicates only pairs of model locations with connectivity.

#### Application of non-pharmaceutical interventions

In the absence of vaccines and other preventive treatments, non-pharmaceutical interventions, such as school closures, social distancing, stay-at-home directives, and testing and isolation are critical strategies for reducing disease transmission. The model enables users to specify non-pharmaceutical interventions as changes to the basic reproductive number (*R*_0_) and the inverse of the infectious period (7), for pre-specified periods of time and to apply non-pharmaceutical interventions to all or subsets of model locations independently. Non-pharmaceutical interventions can be implemented with fixed or distributional effectiveness. In addition, users may specify a rate of fatiguing intervention effectiveness (e.g., declining adherence to a policy) over a certain number of days. This format enables flexibility in scenario planning; for instance, the model can be used to examine the effects of chaining multiple interventions together over time (e.g., school closure then stay-at-home), gradual declining adherence of the population to an intervention, switching interventions on and off over time, and spatially heterogeneous interventions (Figure 2).

**Figure 2:**
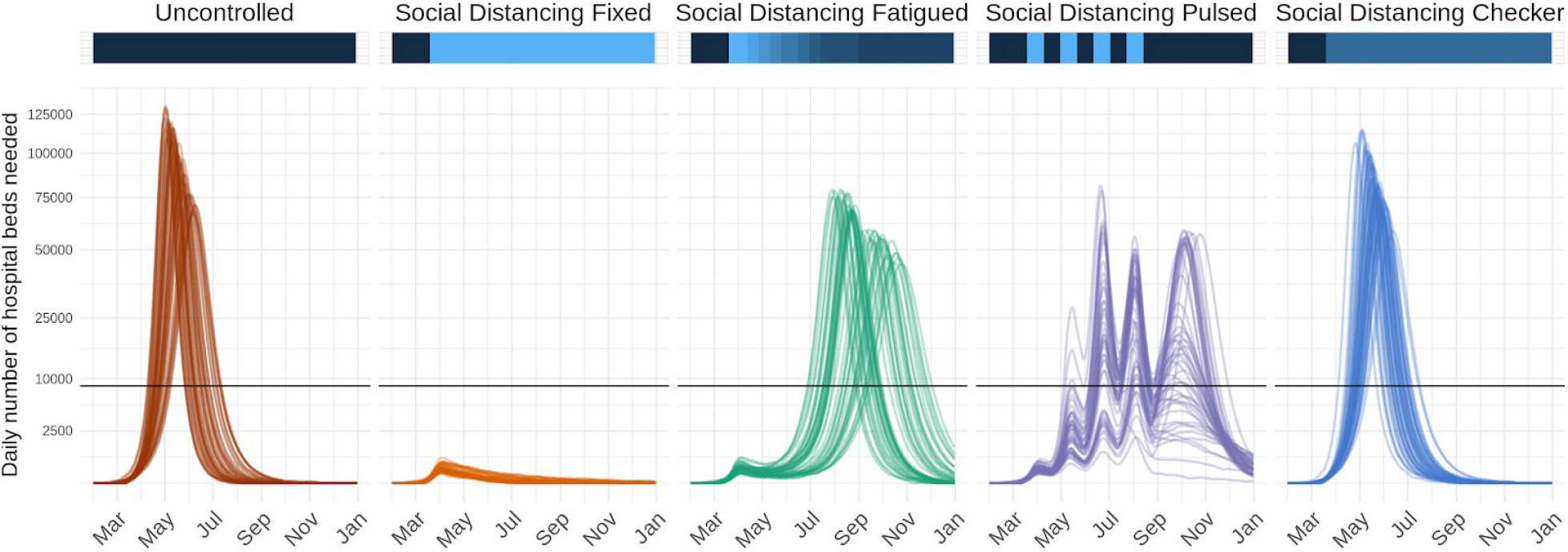
Time series of daily number of hospital beds needed across five possible intervention scenarios in a fictional location with nine counties. Lines represent results from 50 stochastic model simulations. Horizontal black lines represent the total hospital bed capacity in the fictional location, assumed to be n=8661 (3 beds per 1,000 population). The colored horizontal bars along the top visualize the effectiveness of interventions at a given time point along a dark blue to light blue spectrum; dark blue indicates a period with no reductions to transmission, while light blue indicates a period with more restrictive action (i.e., low transmissibility). In the fifth scenario, “Social Distancing Checker,” only 3 of 9 counties implement any non-pharmaceutical interventions, thus differentiating it from “Social Distancing Fixed”.

The effectiveness of a given non-pharmaceutical intervention modulates the daily transmission term below:>

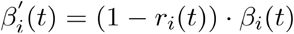

where 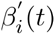 is the daily transmission rate after accounting for intervention effectiveness *r_i_*(*t*) at the specified location *i* at time *t*. When non-pharmaceutical interventions are in effect 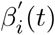 replaces AW in the force of infection term *FOI_i_*(*t*)

Intervention specification is completely user-specified. We include a set of common non-pharmaceutical intervention scenarios that can be applied for user-specified dates and locations (Table S1), which have been compiled according to our review of the literature on the potential impact of non-pharmaceutical interventions on respiratory virus transmission.

### Module 3: Calculation of health outcomes

This pipeline module translates outputs from the transmission model into health outcomes such as hospitalizations and deaths. It takes in counts of daily incident infections and produces daily counts for specific health outcomes at appropriate time delays.

The current default implementation produces hospital and intensive care unit (ICU) admissions, current hospital and ICU occupancy, ventilators needs, and number of deaths. Our modeling of health outcomes assumes that there is some transition probability from infection to death, infection to hospitalization, hospitalization to ICU admission, and ICU admission to ventilator use. In our modeling of health outcomes, we consider the probability of its occurrence (e.g., probability that infections are hospitalized), the time delay relative to its disease course (e.g., time between hospital admission and ICU admission), and where applicable, the duration in a given state (e.g., how long a patient remains ventilated).

The user may specify health outcome probabilities and delays, conditional on the flows described above. For use as default values, we provide tables of parameter values which were derived from a literature review of COVID-19 health outcomes (Table S2).

The pipeline currently contains two versions of this module with different approaches to the specification of health outcome risks: 1) unadjusted, uniform risk and 2) location-specific risks, adjusted by key demographic or health factors in each location.

#### Unadjusted, population-wide health outcome risks

This option generates health outcome estimates with unadjusted risk across all locations, assuming fixed values for all health outcome probabilities, delays and durations.

We assumed that the number of infections admitted to the hospital is a draw from the Binomial distribution, lagged by a fixed time from symptom onset to hospital admission:

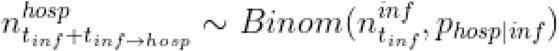

where *n^hosp^* is the number of hospital admissions, *n^inf^* is the number of infections, *t_inf_* is the time of infection, *U_inf→hosp_* is the mean time delay between infection and hospital admission, and *P_hosp|inf_* is the probability of hospitalization given infection (Table 1).

We make similar assumptions for the transitions between other outcomes (hospitalization admission to ICU admission, ICU admission to ventilator use, infection to death):

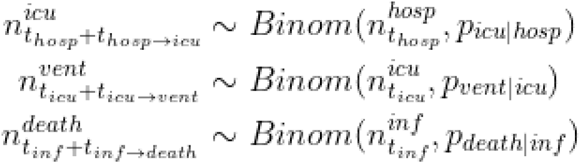

The number of patients currently hospitalized, admitted to ICU, and ventilated are generated from the incident number of events and fixed (non-distributional) user-defined durations for each event.

As information about the death and hospitalization rates were scarce early on in the pandemic, decision makers wanted to consider how things would unfold over different scenarios of health burden. To facilitate these needs, our pipeline separates hospitalization and death rates from the other outcomes, and allows users to consider multiple scenarios for different rates.

#### Location-specific health outcome risks

The health outcomes of SARS-CoV-2 infection can vary greatly between locations due to differences in age distributions and health status between populations. For this reason, the hospitalization module also supports the specification of location-specific relative risks. Users may provide a wide format data file with standardized variable names for the transition probabilities and relative risks (columns) by geoid (rows). These transition probabilities are conditional on previous states (e.g., probability of hospitalization given infection is named p_hosp_inf). The probability of hospitalization and death given infection are specified as relative values compared to population-wide averages specified in the configuration file. We note that the location-specific standardizations apply only to the health outcomes, making a critical assumption that all individuals are at equal risk of infection.

While location-specific data is not required to differ for all health outcome transition probabilities (i.e., some probabilities can be constant across all locations), the location-specific file should include the variables described in Table 1 and population-wide average values for the probability of hospitalization (p_hosp_inf) and death (p_death_inf) given infection.

**Table 1.** Health outcome risk parameters

| Column Name (notation, as appropriate) | Description |
| --- | --- |
| `p_hosp_inf` ($p_{hosp\vert inf}$) | probability of hospitalization among infected individuals |
| `p_icu_hosp` ($p_{icu\vert hosp}$) | probability of ICU admission among hospitalized individuals |
| $p_{\text{vent\_icu}} (P_{\text{icu}\vert \text{hosp}})$ | probability of ventilation among individuals in the ICU |
| $p_{\text{death\_inf}} (P_{\text{death}\vert \text{inf}})$ | probability of death among infected individuals |
| $rr_{\text{hosp\_inf}}$ | relative risk of hospitalization (given infection) relative to the average across all geoids |
| $rr_{\text{death\_inf}}$ | relative risk of death (given infection) relative to the average across all geoids |

To facilitate construction of such files, we have created a companion package `covidSeverity` that produces location-specific relative death and hospitalization rates based on the age distribution of the local population [18]. The package generates these outputs for US counties based on data from the US Census Bureau, and we provide this as a model input file as part of the main pipeline implementation (see COVIDScenarioPipeline/sample_data/geoid-params.csv). The package also includes built-in functionality to pull data from WorldPop [19] and generate adjustments for any location of interest [18].

The `covidseverity` package applies a logistic generalized additive model (GAM) with a penalized cubic spline for age and random effect for age-specific estimates of risk of each health outcome from the literature, thus producing estimates of risk for 10-year, aggregated age categories. These age-specific estimates are then applied to the population age distribution in a given location.

### Summarization of model outputs

This component of the pipeline provides wrapper functions for the lightweight summarization of model outputs into quantiles, plotting functions for common figures, and R Markdown templates to facilitate the rapid generation of technical reports. This module is available in the R package `report.generation` in the Github repository “HopkinsIDD/COVIDScenarioPipeline.”

We provide two key functions that read and process individual transmission (`load scenario sims filtered`) and health outcome (`load hosp sims filtered`) model output files. In managing individual files with these functions, we reduce the processing time and memory load. Both of these functions take processing functions as arguments, thus enabling aggregation and filtering to occur at the level of individual files.

The package contains technical report R Markdown templates for US states, US counties, and countries, a diagnostic report template called “sanity_check_report,” and a template that is maintained solely for integration testing. When `report.generation` is installed and loaded, the templates become available to the user. Many parameters are drawn from the configuration file automatically and pre-written R Markdown chunks about the module options and methods can be referenced within the package.

Common figures include summary tables and time courses for estimated health outcomes under different interventions, maps that portray cumulative cases at the county level, comparisons between model estimates and observed cases and deaths, and visualizations for when ICU and ventilator capacity for each county is exceeded. The vignette below walks through example report outputs in more detail and we provide a template-generated report example in the Supplementary Material (Example Report).

### Model specification

All components and settings for simulations from the COVID Scenario Pipeline model are specified in an easily-modifiable YAML configuration file. We describe different options in detail in the Results and an example of the complete set of configuration options is provided in the Supplementary Material.

### Model access and use

The project is open-source under the GNU General Public License v3.0 license, and code is available at https://aithub.com/HopkinsIDD/COVIDScenarioPipeline. The master branch of this repository consists of a Python package “SEIR” and two R packages “hospitalization” and “report.generation,” which correspond to the second, third, and fourth modules of the model pipeline (https://zenodo.org/badae/latestdoi/245866576y Air importation-based seeding is implemented in the `covidImportation` package (https://aithub.com/HopkinsIDD/covidlmportationy while seeding according to the earliest identified cases is performed in scripts within the “HopkinsIDD/COVIDScenarioPipeline” repository.

## Results: scenario modeling vignette

Here, we present a vignette with 50 model simulations in a fictional setting (Location X) with nine counties (named A through I) in demonstrative intervention scenarios from January 31 to December 31, 2020. In this vignette, we walk through how to set up and run the pipeline, demonstrate some of the spatial and temporal features for modeling non-pharmaceutical interventions, and display some of the plotting functions useful for summarizing the model output.

To run the model, users will require the functionality of the “HopkinsIDD/COVIDScenarioPipeline” repository and a second GitHub repository specific to their model location. A template for such a spatial repository may be found at “HopkinsIDD/COVID19_Minimal” (https://aithub.com/HopkinsIDD/COVID19_Miniman), and complete details on downloading and running the model are available in the template’s wiki at https://aithub.com/HopkinsIDD/COVID19_Minimal/wiki.

Once we have a working environment, the next step is to create a configuration file to describe the model specifications. A skeleton configuration file is included in the C0VID19_Minimal template, and the full configuration file accompanying the results of this vignette is in the Supplementary Material.

First, we specify the broad parameters of the run, such as the date range covered and the number of simulations to run (see first section in Supplementary Material, Example YAML Configuration File).

Next we provide spatial setup information identifying the location of files containing geographic data and the geographic targets for modeling. The `spatial_setup` section of the configuration file identifies a geographical data (geodata) file that contains the population data for each county or administrative subunit in the location(s) of interest and a mobility matrix file that contains the daily trip counts for each pair of counties. Users may employ the R scripts “build_US_setup.R” or “build_nonUS_setup.R,” which are provided with the repository, to generate compatible geodata and mobility matrix files. For users modeling non-US settings, the Github repository “COVID-19-Mobility-Data-Network/mobility” can be used to fit real-time or sparse travel data with mobility models in order to generate similarly compatible mobility matrixes [20,21].

We note that in situations with high cross-border mobility, it may be important to model a region larger than the location of interest in order to appropriately capture disease transmission risk in a given location. Here, we model Locations X, Y, and Z even though Location X is the sole location of interest:

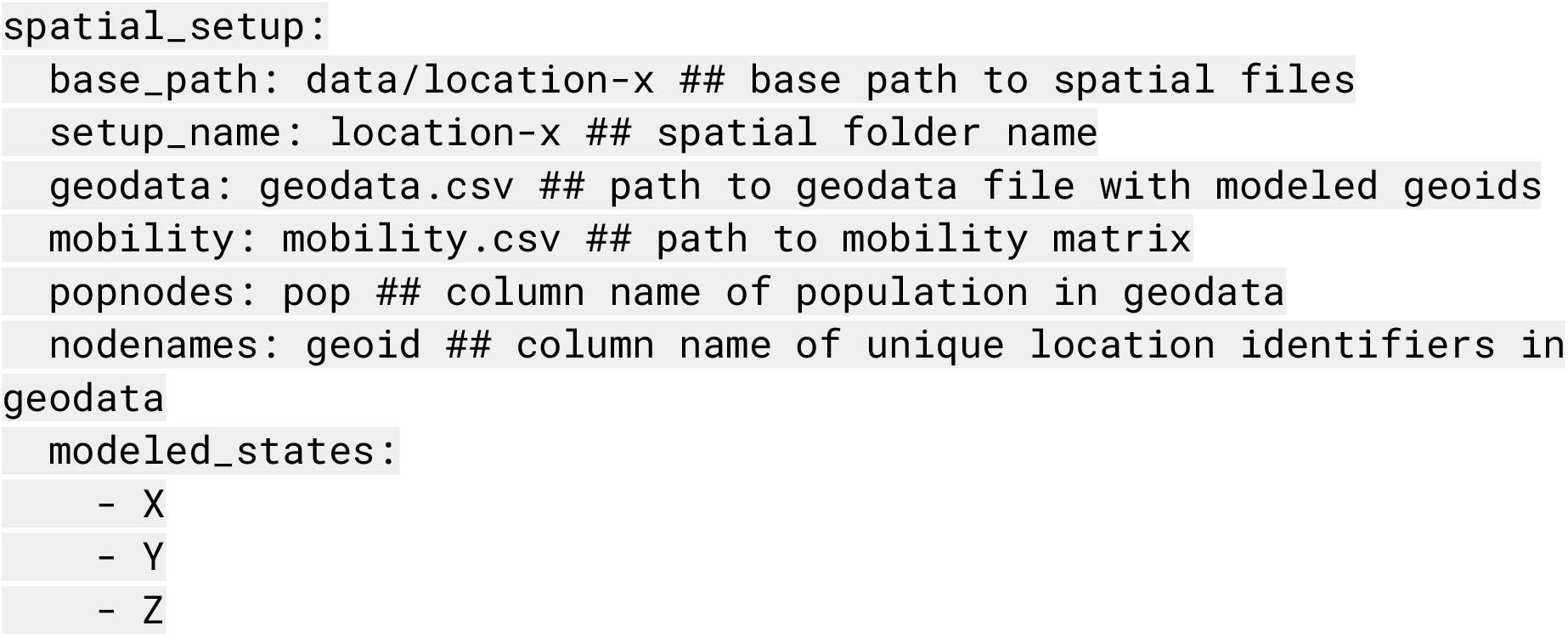

We then specify the locations of the seeding files (see `seeding section` in Supplementary Material, Example YAML Configuration File), with a separate section to describe the air importation model parameters as appropriate (see `importation` in Supplementary Material, Example YAML Configuration File).

Next, we specify the parameters that determine the course of the disease. The `seir` section of the configuration file defines parameters used in the SEIR disease transmission model, including the level of population mixing (where 1 is homogeneous mixing, and < 1 is heterogeneous), the incubation period of the virus, the infectious period, and the baseline basic reproductive number *R*_0_. These values may be fixed or drawn randomly from a distribution, according to the configuration file:

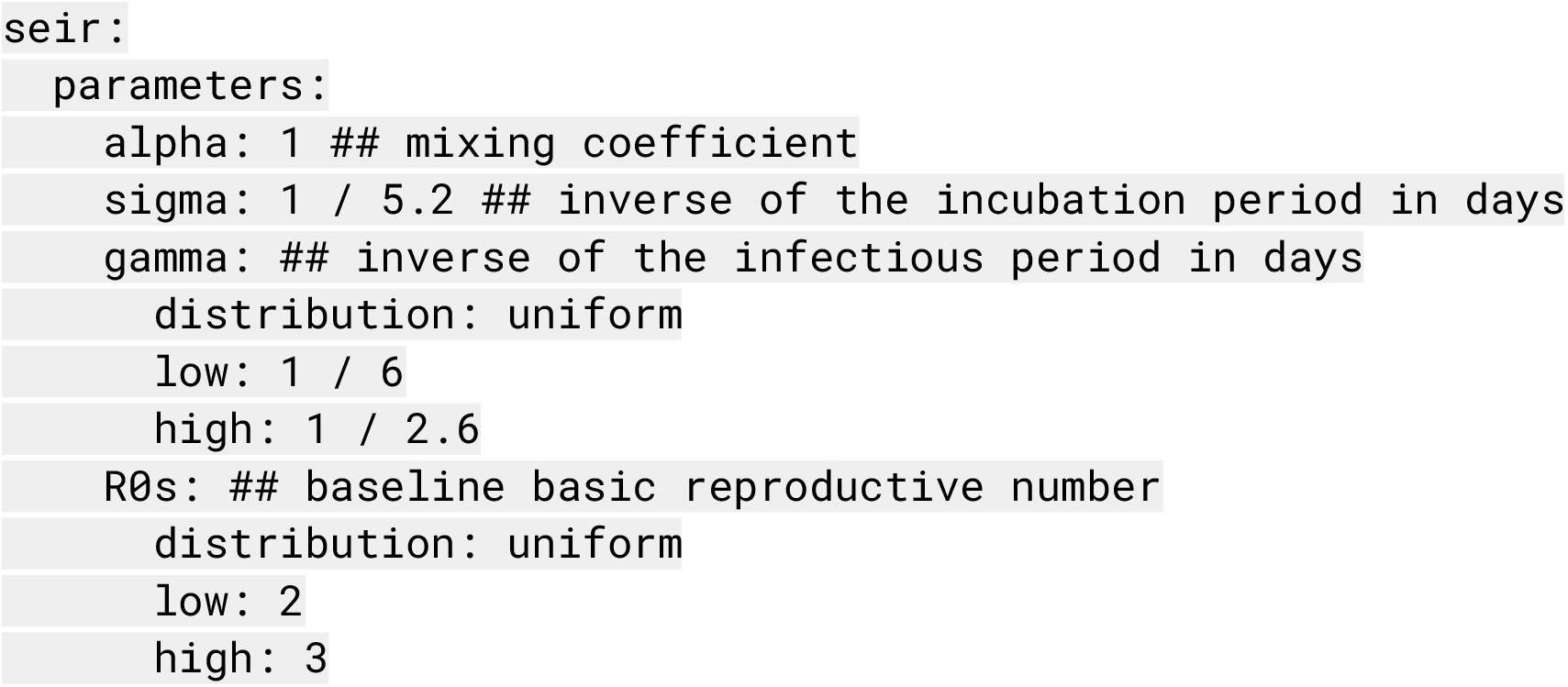

We typically parameterize sigma and gamma in our model with estimates of the range of the serial interval (SI) or generation time, such that

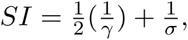

which assumes that the average infection occurs halfway through an index case’s infectious period.

The next step is defining the modeled intervention scenarios. Here, we considered five scenarios in our vignette example: 1) a no intervention scenario (named `Uncontrolled`), in which *R*_0_ remains unchanged over the course of the outbreak; 2) social distancing measures with fixed effectiveness in place from March 19 to December 31 (`SocialDistancing_fixed`); 3) social distancing measures with declining compliance, which were modeled as 10% reductions in effectiveness every 2 weeks beginning March 19 (`SocialDistancing_fatigued`); 4) social distancing measures following a 3-week on-off “pulsing” cycle from March 19 to August 12 (`SocialDistancing pulsed`); and 5) social distancing measures with spatial heterogeneity (`sociaiDistancing_checker`), where three of nine counties implement social distancing measures with fixed effectiveness from March 19 to December 31.

An intervention may be specified in a single block for all model locations, as in the case of the `socialDistancing_fixed` scenario, or for unique location identifiers (“geoids”) as in the

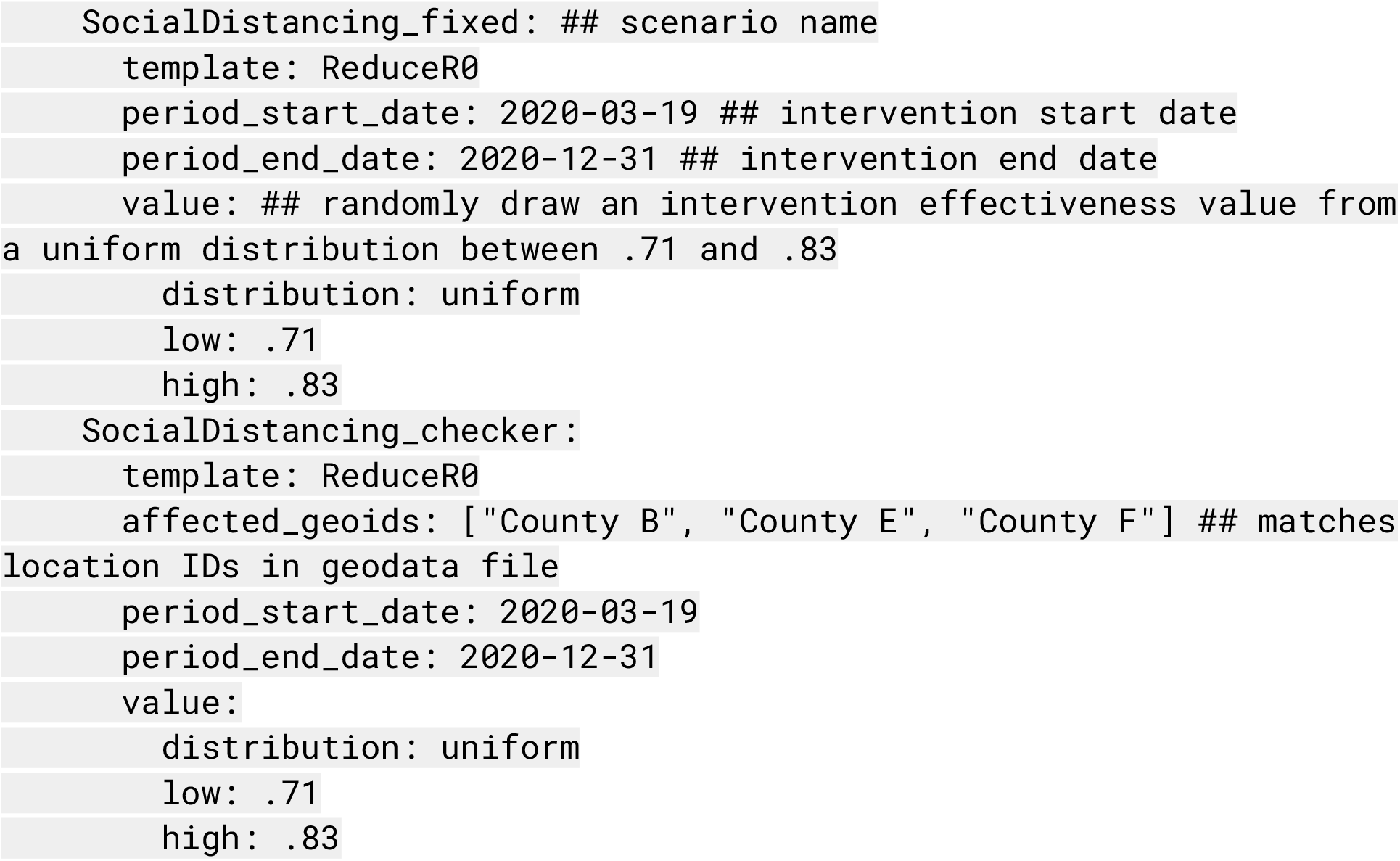

Other intervention scenarios require multiple blocks to be stacked together, as in the case of the `socialDistancing_pulsed` scenario, shown in truncated form below:

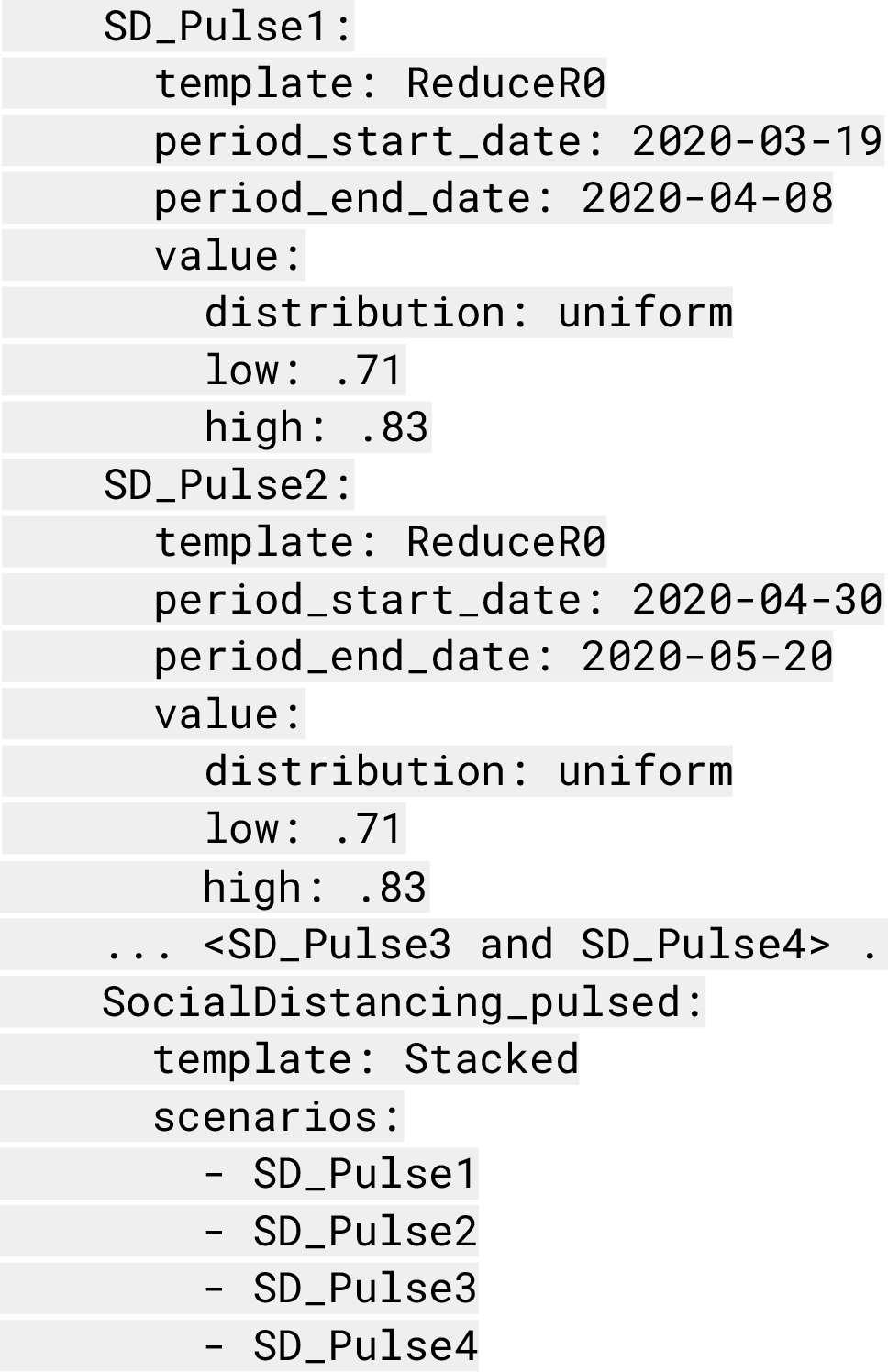

Then, we set up the health outcome risk specifications. In the `hospitalization` section of the configuration file, we specify whether the model calculates health outcome risks with age-adjusted estimates, the average infection fatality ratios (IFR), and the time delays between different health outcomes. The time delays are modeled with lognormal distributions and parameterized with the log median and log standard deviation. For ease of use, we provide a table of estimates found in the literature in Table S2.

After the simulation runs complete, we can produce rapid summaries of the results using R Markdown templates provided by the `report.generation` package. The `report` section of the configuration file specifies scenario labels and colors, IFR scenario labels, and table display dates. These settings, along with other model parameters, can be loaded into a technical report template.

Here we describe how the `state_report` template in the `report.generation` package can be used to display our model results as an example. The full template-generated report linked to this vignette is provided in the Supplementary Material (Example Report). First, the configuration file is loaded to pull in the model settings and file paths. Then, we use a number of predefined functions to load and plot the data.

For example, one standard report figure compares time series of the daily number of hospital beds needed across intervention scenarios (Figure 2) using the `load_hosp_geocombined_totals` and `plot_ts_hosp_state_sample` functions from the `report.generation` package in order to load and plot the data. To plot variations of these figures, we only need to change which health outcome variable is specified in `plot_ts_hosp_state_sample`.

While Figure 2 displays aggregate results, our reports also provide location-specific risk and logistical outputs at the county- or administrative subunit-level. For example, we present the age-adjusted infection fatality ratios and risk of ICU admission among hospitalized infections for the modeled counties within the distribution of all counties in the United States in Figure 3A and Figure 3B. Using the `load hosp geounit relative to threshold` and `plot needs relative to threshold heatmap` functions from `report.generation`, we display the potential need for beds in excess of health system capacity by model location in our reports (Figure 3C). Values of the location-specific healthcare capacity, represented by the number of staffed hospital, acute care, or intensive care beds available and/or number of ventilators available, are user-defined; these can be based on assumptions of the average availability per person or input from data where available.

A map of model outputs makes it easier to visualize spatial and temporal heterogeneity in different intervention scenarios (Figure 4). We provide this functionality in the `load cum inf geounit dates` and `plot geounit map` functions in the `report.generation` package.

**Figure 3:**
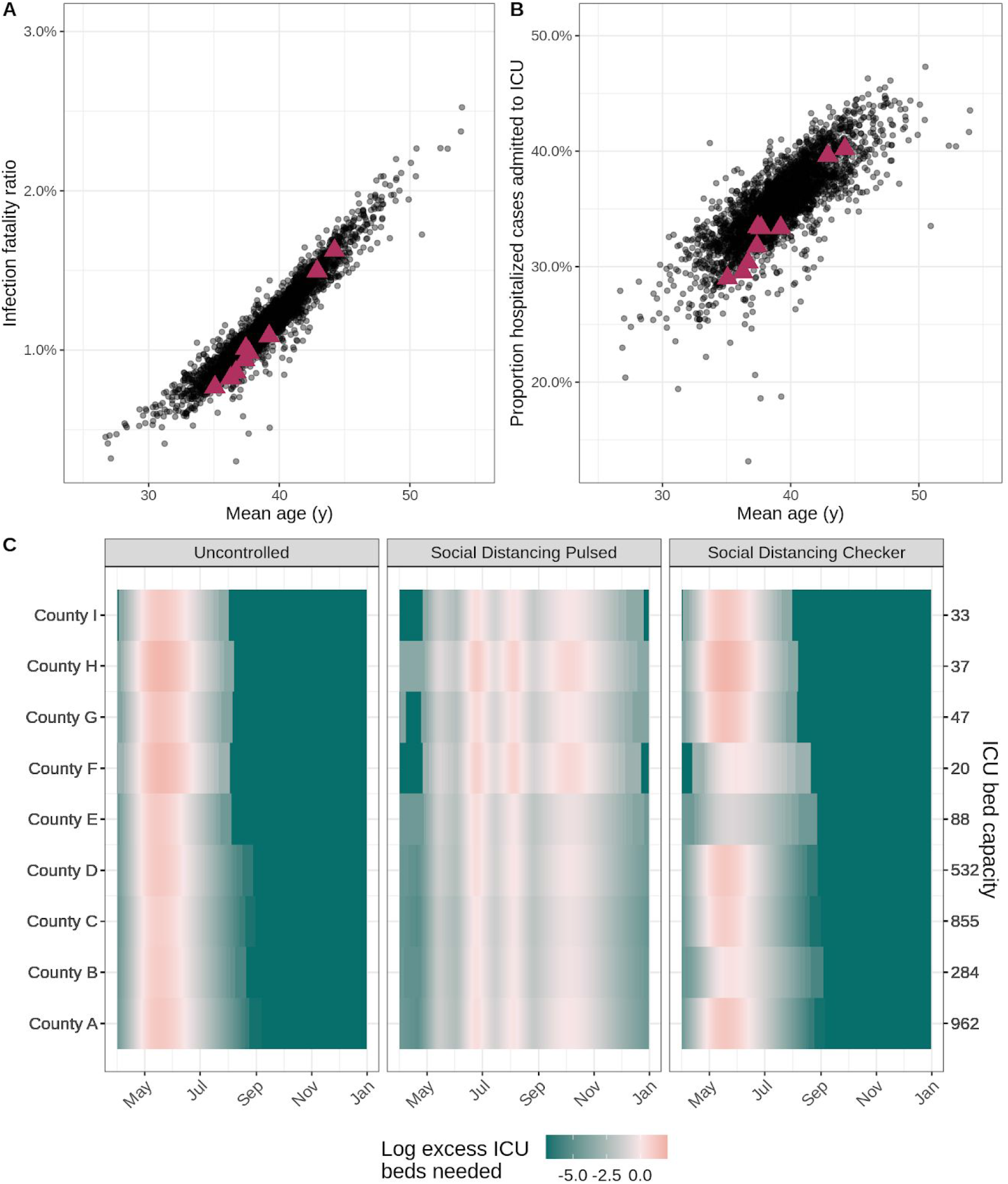
Health outcome risks and logistical needs for fictional counties. In the scatterplots, each point indicates (A) the age-adjusted infection fatality ratio and (B) risk of ICU admission given hospitalization by mean age for a county within the United States. Data for the nine fictional counties in our vignette is marked by magenta triangles. (C) The heat maps display county-level ICU bed needs, shaded according to the log ratio above or below the assumed ICU bed capacity (secondary y-axis) in each county (primary y-axis) for three example intervention scenarios (panels). The salmon pink shading indicates periods of time where ICU bed needs exceed capacity in the fictional counties.

**Figure 4:**
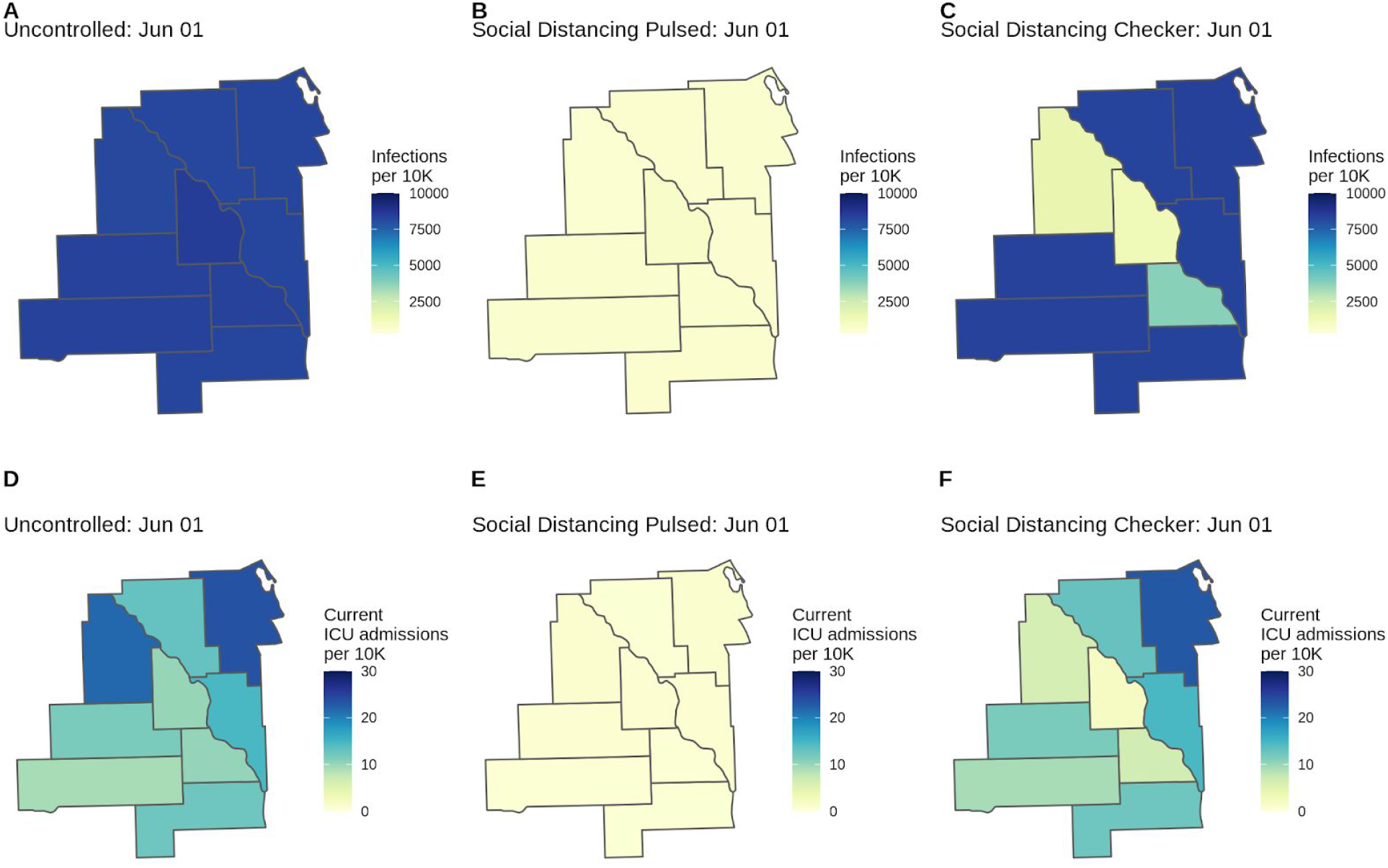
County-level COVID-19 risk for three scenarios (Uncontrolled, Pulsed, Checker) in the fictional Location X. Choropleths for model outcomes on June 1, 2020 of the (A-C) cumulative infection rate per 10,000 population and (D-F) number of patients currently admitted to the ICU per 10,000 population for the Uncontrolled, Social Distancing Pulsed, and Social Distancing Checker intervention scenarios. County-level variation in attack rates can arise from differences in risk of importation, mobility patterns connecting subdivisions, and differences in non-pharmaceutical interventions applied in each location. If location-specific health outcome risks are specified (as are age-standardized health outcome risks in this example), this may serve as another source of county-level variation.

Each report template is equipped to load static R Markdown reference chunks, which we have pre-written and provided with the package. These chunks provide details on our methods, limitations, and key references, pulling in parameters from the configuration file as needed.

## Discussion

We present our scenario pipeline as an open-source modeling framework that aims to balance epidemiological rigor with the flexibility and urgency required by public health policymaking. The modularity of our framework has enabled us to adapt our assumptions about COVID-19 epidemiology, transmission, and health outcome risks in response to emerging information and to different settings. The pipeline implementation of non-pharmaceutical interventions is highly adaptable for policymakers desiring to compare the impact of different potential scenarios.

Throughout the course of the pandemic, we have adapted the default settings of our pipeline in response to the changing needs and questions of our collaborators. At the beginning of the pandemic, air importation seeding was a critical determinant of epidemic onset in specific locations. Now that cases of COVID-19 are present worldwide, we have shifted towards developing more empirical methods of epidemic seeding that better match trajectories of confirmed cases in specific locations as policy questions have shifted to more operational needs. Further, as new data emerged, we moved from calculating unadjusted health outcomes to health outcomes based on age-standardized risk of hospitalization, ICU admission, and death according to emerging case-study data.

As the COVID-19 pandemic continues, we plan to continue expanding the scope of the COVID Scenario Pipeline to changing needs and questions. Future model releases will include a health outcomes model expansion that will enable a multiplicity of pathways to ICU occupancy, ventilator usage, and death. As questions have shifted to near-term operational needs, we have also begun to incorporate inference into our models, thus enabling the calibration of model trajectories to deaths and confirmed case counts, short-term forecast of health outcomes, and estimation of location-specific transmission parameters and NPI effectiveness.

The current implementation of our model has several limitations. We do not explicitly model the role of asymptomatic transmission or other factors that may lead to biases in reporting, and we assume that only one progression in health outcome severity exists (infections to hospitalization to ICU to ventilator use), although we know that many disease course progressions are possible. In addition, the delays and durations involved in our health outcomes progression are fixed values, despite high variability in the estimates of these values. Our epidemic simulations do not account for age-specific transmission, so our model cannot capture the impact of strategies such as cocooning of high-risk age groups beyond population-level reductions in disease transmission. However, the modular approach taken is meant to allow for easy substitution of models with improvement in any of these areas while still taking advantage of other pipeline components. This flexibility does come at a cost, as the modular pipeline approach requires us to write and read files at the end and beginning of each phase, respectively. This procedure requires more disk space and input/output steps than other modeling approaches that can hold all of the necessary data in memory until a single output is produced at the end. Still, these slowdowns are not critically limiting; we have been able to run 1000 county-level simulations of the United States in less than 10 minutes on a 96 core server.

These limitations point to a broader need to consider the totality of evidence generated by epidemiologic models. While our approach is well-suited to answering policy questions about interventions, it is critical for policymakers to explore projections from multiple models in order to understand the range of possible trajectories and the sensitivity of results to different assumptions. Models that incorporate individual-level behaviors may be better for considering the impact of specific contact tracing strategies or location-specific measures like workplace occupancy or symptom screening policies [22]; models incorporating real-time mobility data can best characterize the impact of movement-related restrictions [23]; models with age-specific transmission may provide more detail on the impact of age-specific interventions like “cocooning” [24] or closing and opening schools; still other models are particularly suited to address questions about health systems burden and forecast operational needs [5,25]. Integrating knowledge from multiple models, where appropriate, with careful consideration of the assumptions and appropriate applications of each model, will strengthen response and preparedness [26].

Our flexible modeling pipeline brings an important voice to this “conversation” of models, by allowing rapid and flexible specification and simulation of even very complex intervention scenarios, and providing flexibility to rapidly update models as our understanding of a disease changes. This approach only reaches its full potential when parameters are based on careful and ongoing consideration of the literature and available data. But, when appropriately used as part of an iterative approach to decision making, this pipeline can be a valuable tool for public health decision making.

## Data Availability

All model code is publicly available on Github in the repositories HopkinsIDD/COVIDScenarioPipeline, HopkinsIDD/covidImportation, and HopkinsIDD/covidSeverity.

https://github.com/HopkinsIDD/COVIDScenarioPipeline

## Author Contributions Statement

KHG, JK, HRM, SAT, LTK, JL, and ECL wrote the first draft of the manuscript. JCL, JK, KHG, and SAT wrote the core model code. KHG and HRM prepared the manuscript figures. HRM prepared the supplementary material. SS, JW, KK, and JPS contributed to model and code optimization and documentation. JL and ECL guided the design, analysis, and execution of the project. JCL, KHG, JK, HRM, SAT, SAL, LTK, JP-S, JL, and ECL contributed to study design and analysis. All authors contributed model code and reviewed the manuscript.

## Acknowledgments

We acknowledge technical support from Amazon Web Services, with particular thanks to Pierre-Yves Aquilanti, Karthik Raman, Arun Subramaniyan, Greg Thursam, and Anh Tran for their time and expertise.

